# Neuron-Enriched Extracellular Vesicle MicroRNAs Reflect Hormone-Sensitive Neural Pathway Changes from Early to Late Perimenopause

**DOI:** 10.64898/2026.04.24.26351678

**Authors:** Rebekah L. Frye, Dana M. Lapato, Christopher Sikes-Keilp, JoAnn V. Pinkerton, Jennifer L. Payne, Vasily Yakovlev, Roxann Roberson-Nay

## Abstract

The menopausal transition represents a major neuroendocrine shift marked by declining estradiol and progesterone, rising follicle-stimulating hormone, and increased vulnerability to cognitive and affective symptoms. Despite extensive evidence of hormone-related neural changes, few biomarkers directly index hormone-sensitive neuronal adaptations *in vivo*. Neuron-enriched extracellular vesicles (nEVs) isolated from blood provide a minimally invasive window into central nervous system (CNS) biology by carrying microRNAs (miRNAs) linked to neuronal regulatory processes. This pilot study tested whether L1 cell adhesion molecule (L1CAM)–positive nEV miRNA profiles differ between early (STRAW stage −2; n = 22) and late (STRAW stage −1; n = 24) perimenopause. A pooled discovery screen of 179 miRNAs identified 10 candidates with substantial fold-change differences between groups; these were then quantified at the individual level using qPCR. Linear mixed-effects models showed a significant main effect of STRAW stage, with late perimenopause associated with higher ΔCq values (lower overall expression) across the miRNA panel. The miRNA x STRAW stage interaction was not significant, indicating a coordinated shift across the measured miRNAs rather than miRNA-specific regulation. No evidence of an association between nEV miRNA expression and current estradiol levels or menopausal symptom severity was observed. Bioinformatic analysis of predicted mRNA targets identified significant enrichment of the gonadotropin-releasing hormone (GnRH) receptor pathway, along with related growth factor, immune, and intracellular signaling pathways, with preferential expression in brain-relevant tissues. These findings are consistent with stage-related differences in hormone-sensitive neuronal regulatory processes across the transition.

## Introduction

The menopausal transition is increasingly recognized as a neurobiological inflection point rather than a purely reproductive milestone. Declining estradiol and progesterone and rising follicle-stimulating hormone (FSH) and luteinizing hormone (LH) influence multiple neural systems. Estrogen receptors are highly expressed in hippocampal and prefrontal regions, where they modulate synaptic plasticity, neuroprotection, and metabolism^1–3^. Estradiol withdrawal has been linked to altered dendritic spine density, reduced long-term potentiation, and compromised mitochondrial resilience^4–6^. At the same time, FSH may directly affect neuroinflammatory and degenerative pathways^7^, while progesterone supports myelination and anti-inflammatory signaling^8^. Together, these hormonal fluctuations shape synaptic, immune, and metabolic processes relevant to cognitive and affective function.

Neuroimaging studies corroborate these effects, documenting changes in gray matter volume, functional activation, connectivity, glucose metabolism, and white matter integrity during the menopausal transition^9–12^. In addition to global structural and metabolic changes, symptom-specific neuroimaging studies demonstrate altered hippocampal and parahippocampal activation during memory tasks in women experiencing vasomotor symptoms (VMS), as well as associations between VMS and white matter hyperintensities independent of sleep disturbance^11–14^. Many women report cognitive changes during perimenopause, particularly in verbal memory, attention, and executive function, and these reports are supported by evidence linking objective VMS to poorer memory performance and markers of compromised brain health^14,15–18^. Together, these findings suggest that hormonally driven symptoms such as VMS may index underlying neural vulnerability. Despite growing evidence of hormone- and symptom-linked brain changes, the field still lacks noninvasive, mechanistically informative biomarkers capable of capturing central nervous system adaptations during perimenopause.

Extracellular vesicles (EVs) provide a promising approach to address this gap. EVs are membrane-bound particles released by most cell types and contain selectively packaged proteins, lipids, and nucleic acids that reflect the physiological state of their cells of origin^19–21^. A subset of EVs circulating in blood is enriched for neuronal origin and can cross the blood–brain barrier, offering a minimally invasive window into CNS biology^22,23^. These neuron-enriched extracellular vesicles (nEVs) have been shown to carry pathological proteins and disease-relevant microRNA (miRNA) signatures in neurodegenerative and neurologic conditions, often preceding overt clinical symptoms^24–26^.

In human plasma, nEVs are commonly operationally defined and enriched using immunocapture strategies targeting neuronal surface markers, most frequently L1 cell adhesion molecule (L1CAM/CD171). Although concerns have been raised that circulating L1CAM may exist in soluble form rather than exclusively on EV membranes^27,28^, multiple lines of evidence support the utility of L1CAM-based enrichment for isolating neuronally derived EV populations. Single-vesicle analyses indicate that a substantial proportion of L1CAM-positive particles co-express neuronal markers^29^, and numerous studies, including our prior work, have demonstrated reproducible enrichment of EVs carrying neuron-specific cargo using L1CAM immunoprecipitation^23,30,31^. Accordingly, L1CAM-enriched EVs are widely used as a practical and biologically informative proxy for neuronally derived EVs in peripheral biofluids.

MiRNAs are particularly valuable cargo within nEVs. They are selectively sorted, protected from enzymatic degradation, and capable of regulating large gene networks involved in synaptic signaling, metabolism, and neuroimmune function^20,32,33^. Because nEVs can traverse the blood–brain barrier, their miRNA content offers a minimally invasive index of CNS regulatory states and may participate in bidirectional neuroendocrine communication^34,35,36^.

To date, no study has examined whether nEV microRNA profiles vary across the menopausal transition. This gap is notable given that perimenopause is characterized not only by shifts in ovarian hormone levels, but by prolonged neuroendocrine instability and cumulative neural adaptation to fluctuating estradiol and progesterone signaling^5,37,38^. Many hormone-sensitive neural processes, including synaptic plasticity, neuroimmune regulation, and cellular stress responses, unfold over extended time periods and may not be well captured by single time-point hormone measurements or symptom reports^15,39,40^. nEV miRNAs offer a potential window into these processes. As neuronally derived regulatory molecules that are selectively packaged, stable in circulation, and capable of modulating large gene networks, nEV miRNAs may encode aspects of longer-term neuronal regulatory states rather than acute endocrine fluctuations^4^/24/2026 4:33:00 PM. We therefore hypothesized that distinct stages of the menopausal transition would be associated with differences in nEV miRNA expression, reflecting stage-specific neuronal regulatory states shaped by cumulative exposure to fluctuating gonadal hormone signaling.

## Methods

### Participants

Participants were drawn from the baseline phase of a larger study examining estradiol variability and stress-system functioning during the menopausal transition (R01MH108690). Participants were originally recruited through the University of North Carolina (UNC) at Chapel Hill School of Medicine between January 2017 and March 2020^41^. Of the original study participants, 46 women meeting STRAW+10 criteria^42^ were included in this study: 22 women in early perimenopause (Stage −2), defined by persistent variability in menstrual cycle length, operationalized as repeated ≥ 7-day differences between the number of days separating the onset of consecutive menstrual bleeding episodes, and 24 women in late perimenopause (Stage −1), defined by skipped menstrual cycles resulting in prolonged intervals of ≥ 60 consecutive days without menstrual bleeding. Importantly, menstrual cycle length refers to the interval from the first day of bleeding to the first day of the next bleed; thus, staging reflects irregular timing of menses rather than closely spaced menstrual periods.

Participants completed the Greene Climacteric Scale^43^, a validated self-report measure of menopausal symptom severity that includes psychological, somatic, and vasomotor subscales, with higher scores indicating greater symptom burden. Serum estradiol (E2) concentrations were quantified using liquid chromatography–tandem mass spectrometry (LC-MS/MS). To address right skewness and satisfy model assumptions, E2 values were log-transformed. Only baseline plasma samples collected prior to any hormonal intervention were analyzed. Demographic and participant characteristics are presented in Table 1. Groups were comparable with respect to age, body mass index (BMI), Greene Climacteric Scale scores, and log-transformed E2 concentrations. All participants provided written informed consent, and institutional review board approval included this secondary biomarker analysis.

**Table 1.** Participant Characteristics by STRAW Stage -2 (early perimenopause) and -1 (late perimenopause).

| **Variable** | **Early**<br>**Perimenopause** | **Late**<br>**Perimenopause** | ***p*** | **Entire Sample** |
| --- | --- | --- | --- | --- |
| **Age (years), Mean (SD)** | 48.86 (2.70) | 50.21 (2.73) | 0.101 | 49.57 (2.77) |
| **Race, n (%)** |  |  |  |  |
| White | 18 (81.8%) | 19 (79.2%) |  | 37 (80.4%) |
| Black | 3 (13.6%) | 2 (8.3%) |  | 5 (10.9%) |
| Asian | 0 (0%) | 1 (4.2%) |  | 1 (2.2%) |
| Other / Multiracial | 1 (4.5%) | 3 (12.5%) |  | 4 (8.7%) |
| **Ethnicity, n (%)** |  |  |  |  |
| Latino | 2 (9.1%) | 2 (8.3%) |  | 4 (8.7%) |
| **BMI (kg/m2)** | 26.80 (6.86) | 27.23 (5.22) | 0.810 | 27.02 (6.02) |
| **Estradiol (log-transformed)** | 4.06 (1.12) | 3.64 (1.57) | 0.307 | 3.85 (1.37) |
| **Greene Climacteric Scale, Mean (SD)** | 11.27 (7.61) | 11.33 (6.58) | 0.977 | 11.30 (7.01) |
| Greene Psychological Subscale | 20.50 (5.99) | 20.71 (5.99) | 0.907 | 20.60 (5.92) |
| Greene Somatic Subscale | 9.64 (3.02) | 9.48 (2.44) | 0.850 | 9.56 (2.72) |
| Greene Vasomotor Subscale | 4.18 (1.33) | 4.29 (1.82) | 0.831 | 4.23 (1.57) |
| **Life Events Scale, Mean (SD)** | 1.45 (1.18) | 1.83 (1.52) | 0.355 | 1.65 (1.37) |

### EV Isolation and L1CAM-positive EV Immunoprecipitation

Plasma EVs were isolated following MISEV2023 guidelines^44,45^. Briefly, EVs were isolated from participant plasma samples using qEV size exclusion chromatography (SEC) columns (Izon Science, IC1-35). Participant plasma (0.5 mL) was loaded onto qEV size-exclusion chromatography columns and diluted to 1.5 mL in pre-filtered (0.1 µm) PBS; EV-containing fractions were collected according to the manufacturer’s protocol. L1CAM-positive nEVs were immunoprecipitated with 5 µg/mL of L1CAM/CD171 Monoclonal Biotin Conjugated Antibody (eBio5G3 (5G3), Thermofisher, 13-1719-82) in the presence of a final concentration of 1% SuperBlock Buffer in PBS (Sigma-Aldrich, 37580) for a final volume of ∼1.6 mL solution.

To capture labeled L1CAM-positive nEVs, 40 μL of Pierce Streptavidin Plus UltraLink Resin (#53117, Thermofisher) was added to 1.6 mL of EV-antibody-inhibitor solution and incubated for one hour at 4°C on a rotator. Following centrifugation at 500 x *g*, the supernatant (containing L1CAM-depleted non-nEVs) was removed and saved at -80°C. The L1CAM-positive nEV pellet was washed three times with 1X PBS that had been pre-filtered through a 0.1 µm pore-size filter.

For nanoparticle tracking analysis (NTA), Western blot, and transmission electron microscopy (TEM), precipitated nEVs were eluted by incubation of the Streptavidin beads with 100 μl of Pierce™ IgG Elution Buffer (Thermofisher, 21028) at room temperature with mixing for 5 min.

### Nanoparticle Tracking Analysis

The concentration and size of EVs were measured using ZetaView Nanoparticle Tracking Analysis for plasma, extracted (total) EVs following SEC, and L1CAM-positive EVs following immunoprecipitation and elution from streptavidin resin. All samples were diluted in PBS to a final volume of 2 ml. Ideal measurement concentrations were found by pre-testing the ideal particle per frame value (140–200 particles/frame). The manufacturer’s default software settings for EVs were selected. For each measurement, three cycles were performed by scanning 11 cell positions each and capturing 80 frames per position under the following settings: Focus: autofocus; Camera sensitivity for all samples: 78; Shutter: 100; Scattering Intensity: detected automatically; Cell temperature: 25°C. After capture, the videos were analyzed by the in-built ZetaView Software 8.04.02 SP2 with specific analysis parameters: Maximum area: 1000, Minimum area 5, Minimum brightness: 25. Hardware: embedded laser: 40 mW at 488 nm; camera: CMOS. The number of completed tracks in NTA measurements was always greater than the proposed minimum of 1000 to minimize data skewing based on single large particles.

### Western Blotting

For protein analysis all samples were loaded and separated by SDS-PAGE gel and transferred to nitrocellulose membranes. The membranes were exposed to antibodies at specific dilutions. The primary antibodies used for WB were the following: anti-L1CAM (CD171) (dilution 1:1000, ThermoFisher), anti-CD81 (dilution 1:1000, ThermoFisher), and anti-TSG101 (dilution 1:500, Cell Signaling). Specific protein bands were detected using infrared-emitting conjugated secondary antibody, anti-rabbit DyLight 800 4X PEG Conjugate (dilution 1:10,000, Cell Signaling). WB images were generated and analyzed using the ChemiDoc Infrared Imaging System (Bio-Rad).

### Transmission Electron Microscopy

Isolated EVs were fixed and prepared for the TEM as was previously described^31^. The TEM analysis was carried out by the Microscopy Core at Virginia Commonwealth University.

### RNA Isolation and Pooled Discovery Profiling

L1CAM-positive EVs were lysed on UltraLink Resin beads with TRIzol reagent (Invitrogen, 10296010) via the manufacturer’s recommendation. Chloroform was used to extract total RNA and precipitated with ice-cold ethanol and 5 μL glycogen (Thermofisher, R0551). Total RNA was extracted following the manufacturer’s recommendations using the miRNeasy MicroRNA Extraction Kit (Qiagen, 217204), converted to cDNA using the miRCURY LNA RT Kit (Qiagen, 339340), and then stored at -80°C.

Initially, all cDNA samples were pooled into two groups: Early Perimenopause and Late Perimenopause based on STRAW+10 categories. Relative expression across these two groups was assessed using a standard Serum/Plasma Focus PCR panel (Qiagen, YAHS-106Y) containing 179 LNA miRNA primer sets of miRNAs that have been commonly found in human plasma. The Serum/Plasma Focus miRNA PCR Panels include potential reference genes and probes for evaluating potential sample contamination or damage (e.g., hemolysis). Each PCR panel also contains set of the negative control (H_2_O) and five sets of RNA Spike-In controls to evaluate RNA extraction (Spike-Ins 2-4-5; concentration ratio 1:100:10000) and cDNA synthesis (Spike-In 6) and to perform inter-plate calibration (Spike-In 3). The amplification was performed in a CFX Opus 384 Real-Time qPCR System (Bio-Rad, 12011452). Amplification was conducted with miRCURY LNA SYBR Green PCR Kit (Qiagen, 339347) using the manufacturer’s recommendations. Individual miRNA significantly differently expressed between Early and Late Perimenopause groups were then validated individually for each participant sample present in the pooled groups approach.

### Individual-Level qPCR Quantification

miRNAs identified as significant in the pooled discovery analysis were validated at the individual participant level using miRCURY LNA miRNA PCR Assay primers, following the manufacturer’s instructions (Qiagen, Cat. No. 339306). Quantitative PCR was performed on a Bio-Rad CFX platform, and amplification curves were analyzed using CFX Maestro software (Bio-Rad, v2.3) to determine quantification cycle (Cq) values. Cq values ≥ 40 were classified as not detected and excluded from quantitative analyses.

For quantitative expression analyses, miRNAs were required to be detected (Cq < 40) in both technical replicates; targets detected in only one replicate were treated as not detected for ΔCq calculations. Expression levels were normalized to the reference miRNA miR-151a-5p. This miRNA was detected in all samples without missing values. Expression did not differ between early and late perimenopause (31.7 vs. 31.5 Cq; mean difference = 0.26 cycles; Welch’s p = 0.76), with a negligible effect size (Cohen’s d = 0.09). Overall variability was low (CV = 9%), and NormFinder analysis incorporating intra- and inter-group variance identified miR-151a-5p as a stable candidate. ΔCq values were calculated as the difference between the Cq value of each miRNA and the reference miRNA.

### Statistical Analysis

ΔCq values were analyzed using linear mixed-effects models to account for the clustered structure of the data, with 10 miRNAs measured per participant at a single occasion. MiRNA species were treated as within-participant repeated measures nested within participant. Models included fixed effects for miRNA species, STRAW stage (-2 [early] vs -1 [late] perimenopause), and their interaction to test whether stage-related effects were broadly distributed across miRNAs rather than driven by a specific subset of miRNAs. Random intercepts were specified for participant (ID) to account for within-subject correlation. Because miRNAs differed in measurement variability, models allowed for miRNA-specific residual variances using a diagonal covariance structure for repeated measures within participant. Models were estimated using restricted maximum likelihood (REML) with Satterthwaite degrees of freedom. Estimated marginal means were obtained for STRAW stage and miRNA species; Bonferroni adjustment was applied for pairwise comparisons among miRNAs.

To evaluate whether STRAW stage effects were independent of chronological age, age was standardized and added as a covariate in a secondary analysis. Secondary models also evaluated associations between ΔCq and menopausal symptom severity (Greene Climacteric Scale total score standardized) and log-transformed estradiol standardized. Given the modest sample size, interpretation emphasized effect sizes and confidence intervals alongside statistical tests.

### Bioinformatic Analysis

Canonical and noncanonical messenger RNA (mRNA) targets were identified for each significant miRNA using miRDB^46,47^, an online database that catalogs predicted miRNA targets. The target list was deduplicated and submitted to the PANTHER knowledgebase to perform gene ontology analysis and identify pathways implicated by the significant miRNAs and their target mRNAs. Because ontological and functional analysis results can vary based on the knowledgebase, additional gene ontology analyses were conducted using EnrichR^48–50^. For all functional analyses, Bonferroni correction was applied to all p-values, and significant results were recorded and reported in order.

## Results

NTA confirmed enrichment of small EVs with expected size distributions (average diameter ∼120 nm). Western blots demonstrated robust EV markers (i.e., intraluminal TSG101 and membrane-bound CD81) and concentration of L1CAM signal in the nEV pellet. Immunoprecipitation of nEVs was considered successful by the presence of L1CAM signal in the nEV fraction (pellet) and the depletion of L1CAM signal in the non-EV fraction (supernatant). TEM verified intact vesicles (see Figures 1A–C).

**Figure 1:**
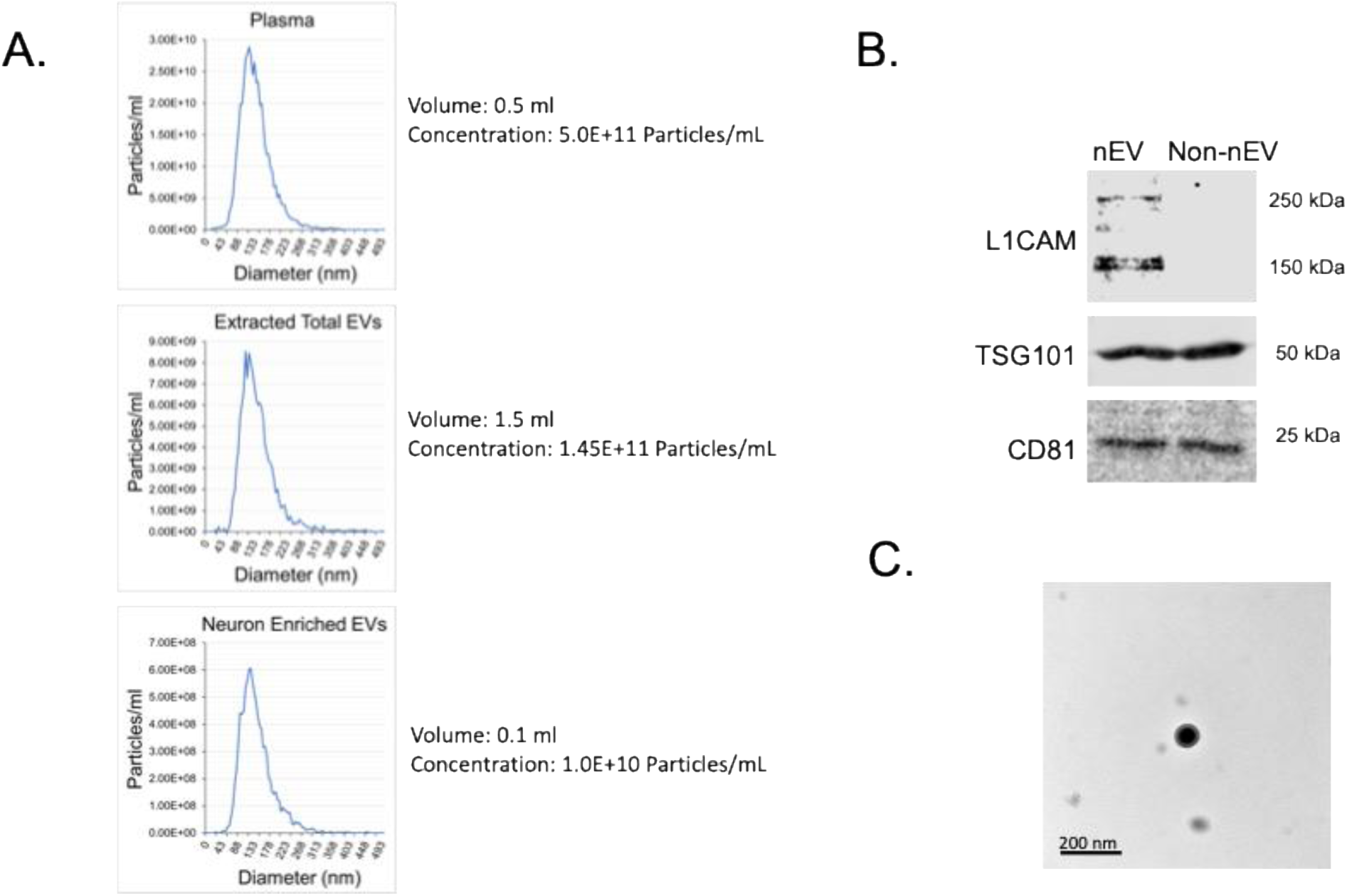
Extraction and characterization of neuron-enriched EVs (nEVs). **(A)** Nanoparticle tracking analysis of EVs in the native plasma, after extraction from plasma by the size-exclusion chromatography (Extracted Total EVs), and after precipitation with Anti-L1CAM(CD171) Ab (nEVs). **(B)** Analysis of EV-specific markers and L1CAM (CD171) protein expression in nEVs and non-neuronal EVs (Non nEVs) (supernatant). Equal amount of total protein from both samples was used for Western blot analysis. Common EV markers (TSG101 and CD81) were present in both nEVs (pellet) and non-nEVs (supernatant), but L1CAM signal was depleted in the supernatant, suggesting expected enrichment of L1CAM-positive nEVs in the pellet. **(C)** Electron microscopy analysis for the nEV fraction showing intact, membrane-bound vesicles.

### STRAW Stage Effects on nEV miRNA Expression

The pooled analysis identified 10 miRNAs showing large fold-change differences between early and late perimenopause (i.e., |log_2_FC| ≥ 5): miR-24-3p, miR-30b-5p, miR-150-5p, miR-152-3p, miR-197-3p, miR-194-5p, miR-425-3p, miR-22-5p, miR-584-5p, and miR-30a-5p (see Figure 2). This pooled analysis was used for candidate miRNA selection. Individual-level qPCR validation demonstrated directionally consistent effects for all ten miRNAs, except for miR-22-5p (see Supplementary Table S1). Across the remaining miRNAs, late perimenopause was consistently associated with higher ΔCq values (i.e., lower expression).

**Figure 2.**
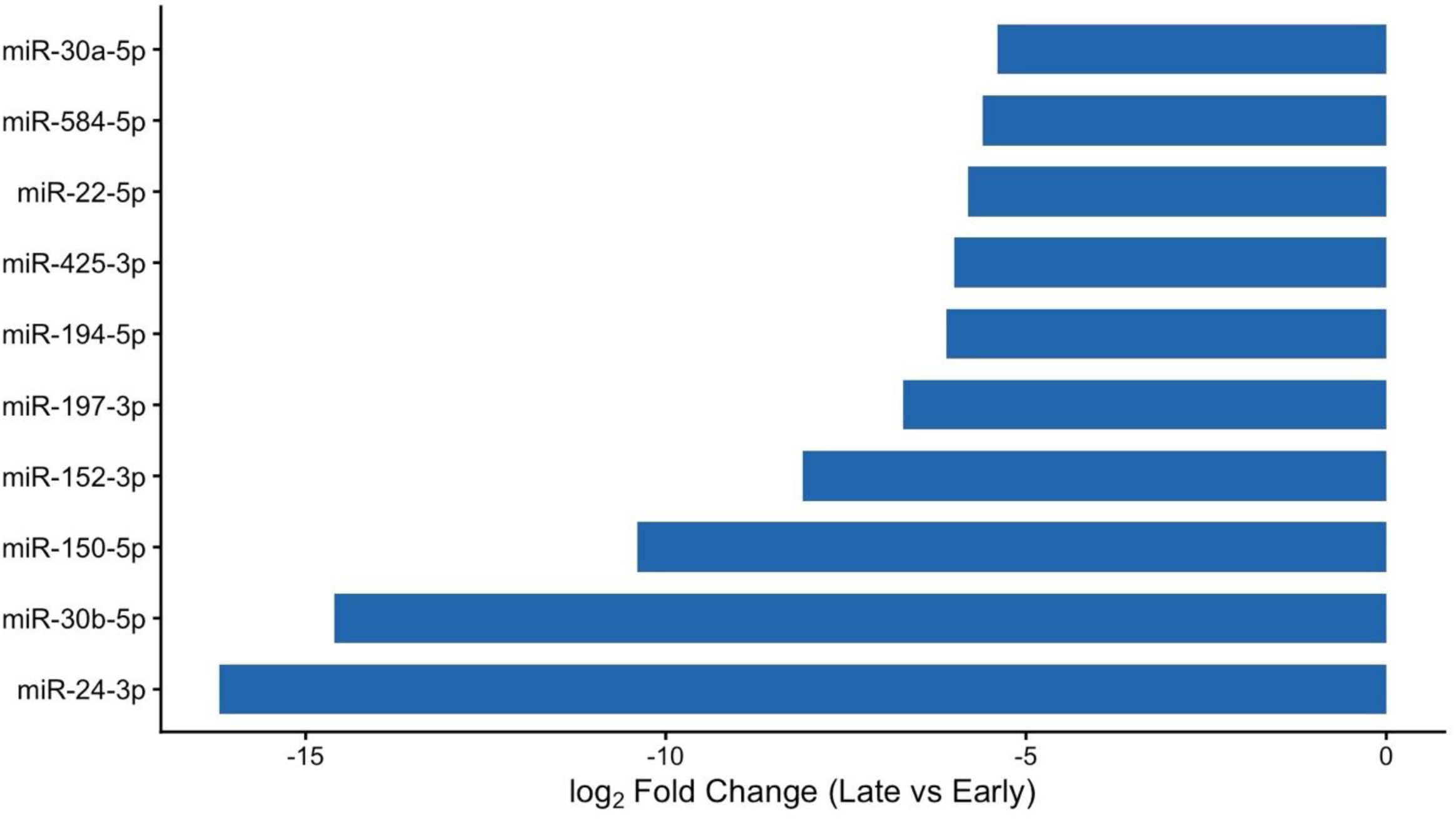
Pooled discovery screen identified miRNAs downregulated in late perimenopause. Horizontal bar plot showing log_2_ fold changes in miRNA expression between late and early perimenopause derived from the pooled discovery screen of neuron-enriched extracellular vesicles. Negative values indicate lower miRNA expression in late perimenopause relative to early perimenopause.

Next, individual-level qPCR data were analyzed using linear mixed-effects models to test the association between STRAW stage and miRNA expression, accounting for participant-level clustering and miRNA-specific residual variability. STRAW stage was significantly associated with ΔCq values (F(1, 57.58) = 5.57, p = .022, partial η^2^ = 0.09), accounting for approximately 9% of the variance in ΔCq values. Estimated marginal means indicated higher ΔCq values in late compared with early perimenopause (early: M = 2.11, SE = 0.16; late: M = 2.62, SE = 0.15), corresponding to lower overall miRNA expression in late perimenopause (see Figure 3). The adjusted mean difference was 0.51 ΔCq units (SE = 0.21), 95% CI [0.08, 0.93] (Bonferroni-adjusted p = .022). This difference corresponded to an estimated 30% lower relative expression in late compared with early perimenopause. See Supplementary Tables S2 and S6 for full results of the linear mixed-effects model.

**Figure 3.**
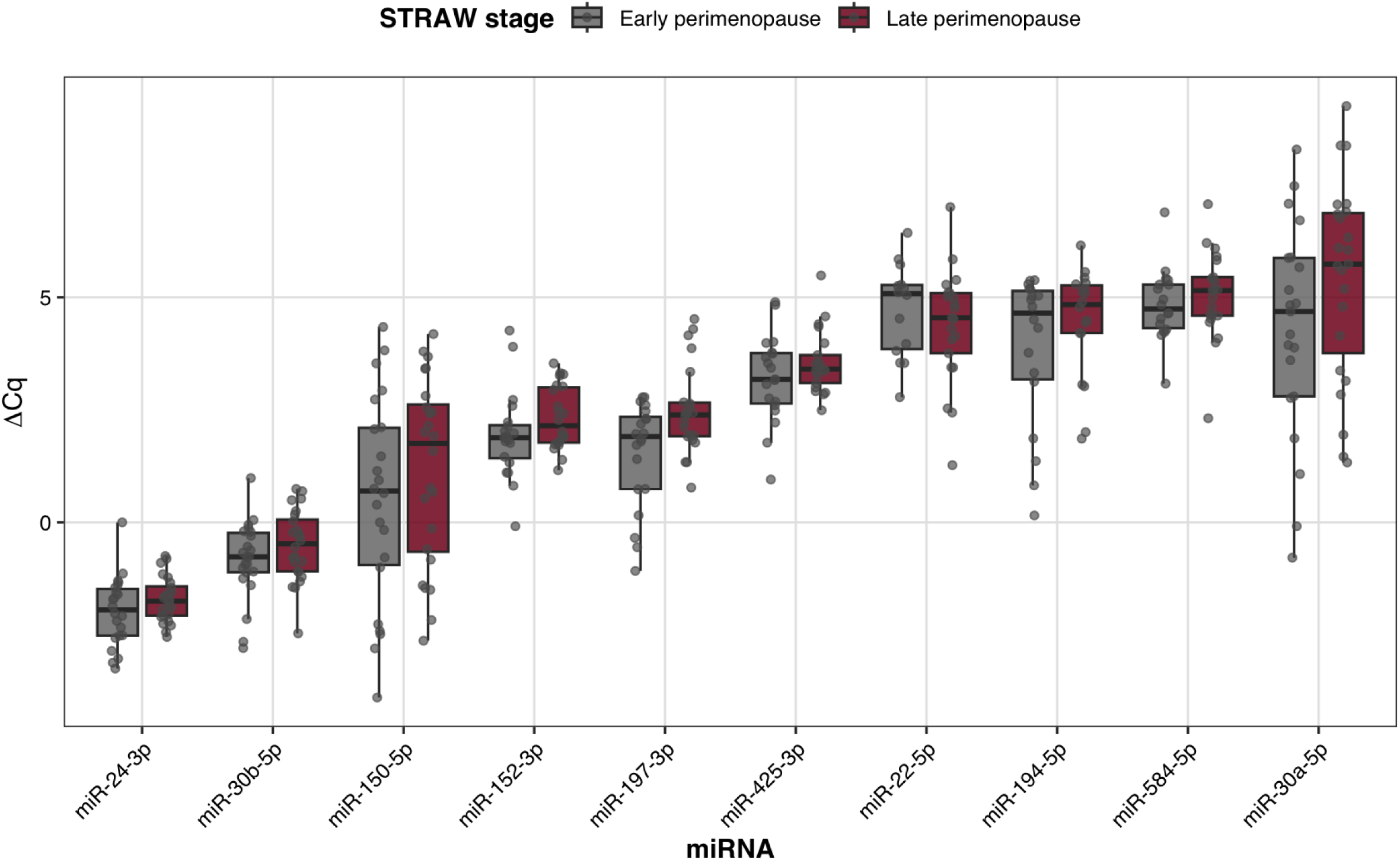
Neuron-enriched extracellular vesicle miRNA expression across the menopausal transition. Box-and-whisker plots depict ΔCq values for ten L1CAM-enriched nEV miRNAs in early and late perimenopause. Boxes represent the interquartile range with the median indicated by the horizontal line; whiskers extend to 1.5x the interquartile range, and points represent individual observations. Higher ΔCq values indicate lower relative miRNA expression. MiRNAs are ordered along the x-axis by baseline expression level. Across miRNAs, late perimenopause is associated with a consistent upward shift in ΔCq values, indicating reduced overall miRNA expression.

**Figure 4.**
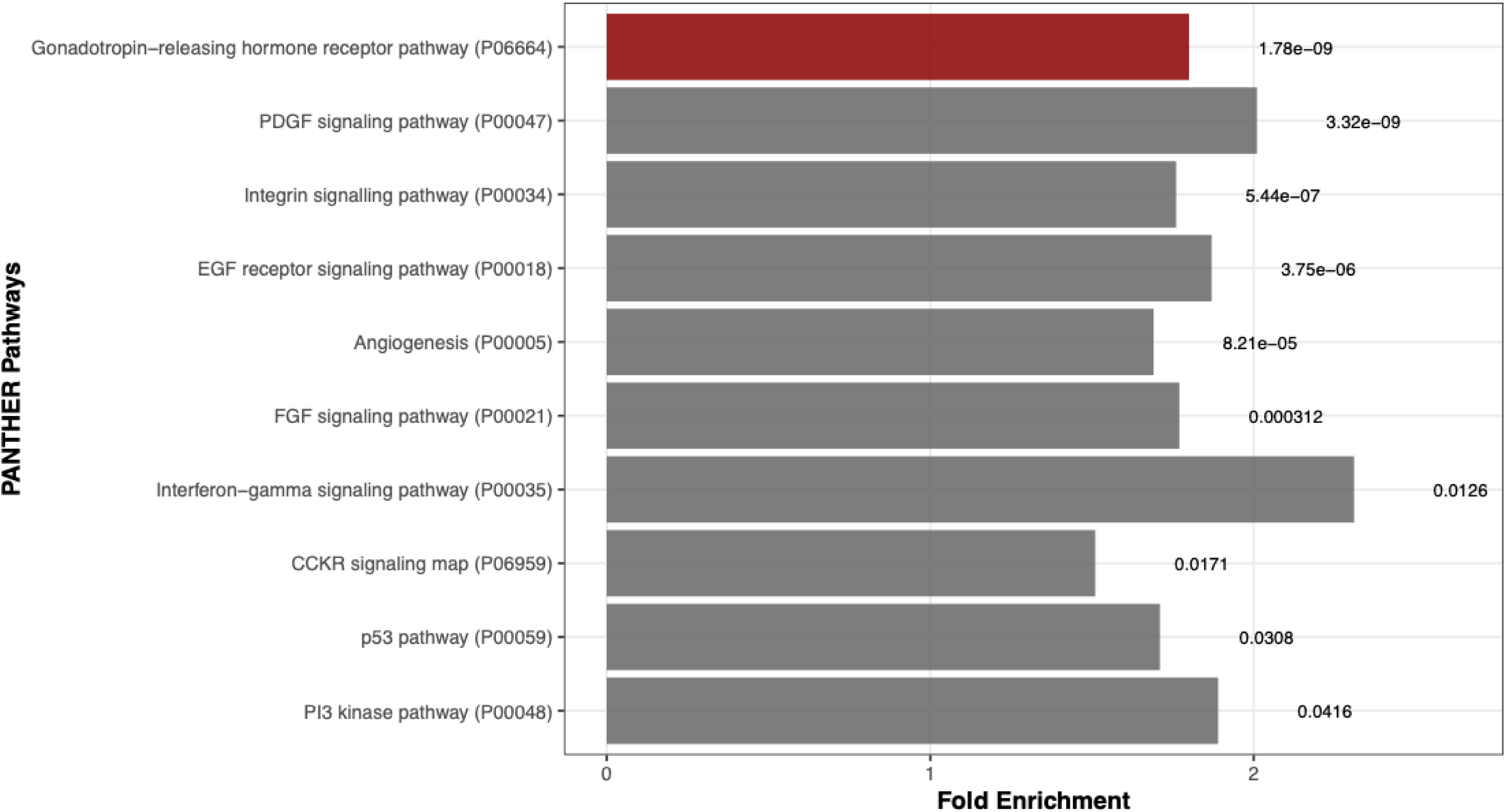
PANTHER pathway enrichment analysis. Horizontal bar plot showing fold enrichment of significantly enriched PANTHER pathways. The top-ranked pathway is highlighted in red; adjusted *p*-values are shown next to each bar.

### miRNA Species and Coordinated Effects

As expected, ΔCq values differed substantially across miRNA species (F(9, 69.48) = 530.77, p < .001), reflecting known differences in baseline expression across the miRNA panel. These differences were confirmed by Bonferroni-adjusted pairwise comparisons (Figure 3) and are reported here to characterize the expression range of the assayed miRNAs. Importantly, the miRNA x STRAW stage interaction was not significant (F(9, 69.48) = 1.27, p = .270), indicating that individual miRNAs did not differ in their stage-related pattern and that the observed effect reflected a coordinated panel-level shift.

### Age-Adjusted Analyses

To determine whether STRAW stage effects were independent of chronological age, age was added as a covariate. The main effect of STRAW stage remained significant after age adjustment (F(1, 54.67) = 5.48, p = .023, partial η^2^ = 0.09), again accounting for approximately 9% of the explainable variance in ΔCq values, whereas age itself explained minimal variance (F(1, 38.37) = 0.08, p = .786, partial η^2^ < 0.01). Estimated marginal means continued to indicate higher ΔCq values in late perimenopause (early: M = 2.10, SE = 0.16; late: M = 2.62, SE = 0.15), with an adjusted mean difference of 0.52 ΔCq units (SE = 0.22), 95% CI [0.08, 0.96] (Bonferroni-adjusted p = .023), corresponding to approximately 30% lower expression in late perimenopause. The miRNA x STRAW stage interaction remained non-significant (F(9, 69.30) = 1.27, p = .270). See Supplementary Tables S3 and S7.

### Menopausal Symptom Severity and nEV miRNA Expression

Menopausal symptom burden, assessed using the Greene Climacteric Scale total score, was not significantly associated with ΔCq values (F(1, 38.86) = 1.60, p = .214), accounting for a small proportion of variance (partial η^2^ = 0.04). The estimated effect was modest (β = 0.13 ΔCq per 1 SD increase in symptom severity, SE = 0.10), with the 95% confidence interval including zero. See Supplementary Tables S4 and S8.

### Estradiol and nEV miRNA Expression

Estradiol concentration was also not associated with ΔCq values (F(1, 38.81) = 0.004, p = .950), explaining negligible variance in ΔCq (partial η^2^ < 0.001). The estimated effect was minimal (β = 0.01 ΔCq per 1 SD increase in log-transformed estradiol, SE = 0.10), with the 95% confidence interval including zero. See Supplementary Table S5 and S9.

### Bioinformatic Analysis

Pathway enrichment analysis was conducted using PANTHER, with Bonferroni correction applied across all tested pathways. Several hormone- and growth factor–related PANTHER pathways showed significant enrichment after correction, including the gonadotropin-releasing hormone (GnRH) receptor pathway (fold enrichment = 1.80, p < .0001), platelet-derived growth factor (PDGF) signaling (fold enrichment = 2.01, p < .0001), epidermal growth factor (EGF) receptor signaling (fold enrichment = 1.87, p < .0001), fibroblast growth factor (FGF) signaling (fold enrichment = 1.77, p < .001), and angiogenesis (fold enrichment = 1.69, p < .001). In addition, integrin signaling was significantly enriched (fold enrichment = 1.76, p < .0001), highlighting pathways involved in cell–matrix interactions and structural signaling. Complete PANTHER pathway enrichment results, including all tested pathways and Bonferroni-adjusted statistics, are provided in Supplementary Table S10.

Immune and intracellular regulatory pathways defined within the PANTHER framework were also overrepresented. Interferon-gamma signaling showed significant enrichment (fold enrichment = 2.31, p = .013), as did the cholecystokinin receptor (CCKR) signaling map (fold enrichment = 1.51, p = .017). Pathways related to cellular stress response and survival, including the p53 pathway (fold enrichment = 1.71, p = .031) and phosphoinositide 3-kinase (PI3K) signaling (fold enrichment = 1.89, p = .042), were likewise significantly enriched. Together, these results indicate involvement of hormone-responsive signaling, growth factor pathways, immune modulation, and intracellular regulatory mechanisms among predicted mRNA targets of perimenopause-associated miRNAs.

Gene Ontology Biological Process (GO BP) enrichment analysis was performed using the same PANTHER framework, with Bonferroni correction applied across approximately 9,300 GO BP terms. 457 GO BP terms survived correction. The GO BP terms with the most significant adjusted p-values were related to regulation of cellular processes (fold enrichment = 1.33, p < 6.4 x 10^-64^), biological processes (fold enrichment = 1.31, p < 1.82 x 10^-62^), and cell communication (fold enrichment = 1.41, p < 1.87 x 10^-50^). GO BP terms with the largest fold enrichment magnitudes included phospholipid dephosphorylation (FE = 2.43), cardiac muscle tissue growth (FE = 2.39), forebrain neuron development (FE = 2.39), and central nervous system neuron axonogenesis (FE = 2.37). See Supplementary Table S11-S13 for Gene Ontology enrichment results, including Biological Process, Cellular Component, and Molecular Function analyses. For completeness, a cross-ontology summary of Gene Ontology enrichment results, spanning Biological Process, Molecular Function, and Cellular Component terms is provided in Supplementary Table S14 and S15.

To provide tissue-level context, predicted mRNA targets were evaluated for tissue expression using data from the Genotype-Tissue Expression (GTEx) Project catalogued through the EnrichR knowledgebase. Target genes demonstrated preferential expression across brain-relevant tissues included in the TISSUES and COMPARTMENTS Curated and Experimental Libraries. Additionally, targets were significantly overrepresented among GTEx Brain genes differentially downregulated with age (GTEx Aging Signatures 2021)^39^.

Exploratory sensitivity analyses were performed by repeating pathway enrichment analyses using two additional annotation frameworks (Reactome, KEGG). Across these analyses, similar core hormone-related and intracellular signaling themes consistently emerged.

## Discussion

In this pilot study, we demonstrate that nEV miRNA expression differs between early and late perimenopause, with late perimenopause characterized by a coordinated reduction in expression across multiple miRNAs. This stage-related effect remained significant after accounting for within-individual correlation across miRNAs, miRNA-specific variance, and chronological age, and was not attributable to any single miRNA species showing a statistically distinct pattern. Together, these findings suggest that nEV miRNAs are consistent with cumulative, hormone-sensitive neuronal adaptation across the transition.

Although individual miRNAs did not show statistically distinct stage-related patterns, the panel included miRNAs previously implicated in neuroinflammatory signaling, synaptic plasticity, and growth factor pathways, including miR-150-5p and members of the miR-30 family^52–55^. These prior associations provide biological context for the observed coordinated panel-level shift.

Within a CNS context, nEV-associated miRNAs reflect neuron-enriched regulatory signaling via extracellular vesicles rather than serving as direct quantitative measures of intracellular miRNA abundance. EV–associated miRNAs are selectively sorted and released in response to cellular state and can reflect alterations in neuronal regulatory tone, metabolic stress, or signaling priorities over extended time periods rather than acute transcriptional changes^56–58^. In human plasma, miRNA cargo measured in L1CAM-enriched EV fractions has been linked to CNS-relevant biological processes and disease-associated signaling changes^23^. The coordinated reduction in nEV miRNA abundance observed in late perimenopause, coupled with the absence of miRNA-specific stage interactions, supports a system-level shift in EV-associated regulatory signaling within this neuron-enriched plasma EV fraction rather than selective dysregulation of individual miRNAs.

Consistent with this system-level pattern, the miRNAs identified in this study are broadly conserved regulatory miRNAs involved in growth factor signaling, immune modulation, and cellular stress responses rather than menopause-specific markers. Such a pattern is consistent with cumulative, hormone-sensitive neuronal adaptation unfolding over extended time periods rather than acute responses to circulating estradiol.

The observed differences in nEV miRNA expression were not associated with estradiol concentrations measured at the time of sampling or with menopausal symptom severity. This absence of a cross-sectional association does not imply a lack of endocrine involvement; rather, it is consistent with evidence that neural and molecular changes across perimenopause reflect longer-timescale responses to variability in ovarian hormone signaling rather than current hormone levels or symptom states. Estradiol levels fluctuate substantially within and across cycles during perimenopause, and single time-point measures are not likely to capture cumulative neurobiological exposure^15,39,17,59^.

These findings also have implications for clinical research and care during the menopausal transition. Current clinical assessment relies largely on symptoms and circulating hormone measurements, which do not capture the underlying neurobiological changes occurring during perimenopause. As a result, clinical interventions are often reactive rather than timed to periods of heightened neural vulnerability or adaptation. The stage-dependent differences in nEV miRNA expression observed here raise the possibility that neuronally derived EV miRNAs may provide a peripheral readout of hormone-sensitive neural regulatory states that are not apparent from endocrine measures alone. Although not currently intended for diagnostic or therapeutic decision-making, such markers could eventually contribute to improved biological staging of the menopausal transition and inform the timing and personalization of hormonal or non-hormonal interventions. Longitudinal studies will be required to determine whether nEV miRNA profiles predict clinical trajectories or treatment response.

Bioinformatic analyses provide additional context for these expression changes. Pathway overrepresentation analysis using PANTHER identified significant enrichment of hormone-responsive and growth factor–mediated signaling pathways among predicted mRNA targets of the validated miRNAs, even after Bonferroni correction. Enriched pathways included gonadotropin-releasing hormone receptor signaling, PDGF, EGF, and FGF signaling, angiogenesis, integrin signaling, and PI3K signaling. These pathways are involved in neuronal plasticity, cellular survival, metabolic regulation, and vascular and extracellular matrix interactions, processes known to be sensitive to ovarian hormone withdrawal and endocrine aging^1,4^.

The enrichment of GnRH receptor signaling is particularly noteworthy in the context of perimenopause, during which alterations in GnRH pulsatility and downstream signaling reflect reorganization of the hypothalamic–pituitary–gonadal axis^37^. Growth factor pathways such as PDGF, EGF, and FGF play key roles in synaptic remodeling, myelination, and neurovascular integrity and have been shown to interact with estrogen signaling in both neuronal and glial cells^1,5^. Similarly, enrichment of integrin signaling and angiogenesis pathways aligns with evidence that estrogen modulates endothelial function and neurovascular coupling, which may change during the menopausal transition^38^.

Immune- and stress-related pathways, including interferon-gamma signaling and p53-related signaling, were also enriched, suggesting potential involvement of neuroimmune modulation and cellular stress responsiveness. Menopause has been associated with shifts in immune tone and inflammatory signaling, and experimental work indicates that estrogen withdrawal can alter glial activation and immune–brain communication^60,61^. While the present study cannot determine whether these pathways are causally engaged, their convergence across analyses supports biological consistency of the miRNA target networks.

Although many Gene Ontology Biological Process terms survived Bonferroni correction, the highest-ranking terms were broad and highly overlapping, reflecting general regulatory, signaling, and developmental processes rather than discrete, pathway-specific mechanisms. This pattern is consistent with the established roles of miRNAs in fine-tuning synaptic signaling, vesicle dynamics, and intracellular regulatory networks rather than acting as on–off switches for single pathways^56^. Cellular Component and Molecular Function analyses further supported involvement of membrane-associated and intracellular signaling machinery, reinforcing the pathway-level findings without introducing independent mechanistic claims.

Tissue-expression analysis using GTEx showed that predicted mRNA targets of the identified miRNAs were preferentially expressed in brain-relevant tissues. This enrichment provides convergent support that the implicated pathways are compatible with CNS-associated biology. When considered alongside prior evidence that nEVs can cross the blood–brain barrier and carry selectively packaged miRNAs linked to neuronal processes, these findings are consistent with, though not definitive proof of, the use of L1CAM-enriched nEV miRNAs as minimally invasive indicators of CNS regulatory activity.

Several limitations warrant consideration. The sample size was modest but comparable to prior nEV studies. Moreover, the pooled discovery approach prioritized large fold changes and may have excluded miRNAs with subtler stage-related differences. Additionally, while L1CAM-based enrichment has been widely used to enrich for neuronally derived EVs, recent work highlights important limitations and ongoing efforts to better define the neuronal specificity and heterogeneity of these EV populations^27,28^. Finally, bioinformatic analyses rely on predicted and curated miRNA–mRNA interactions and should be interpreted as hypothesis-generating rather than definitive evidence of downstream regulation. Future longitudinal studies with repeated EV sampling will be important for determining whether nEV miRNA changes track within-individual neurobiological trajectories across the menopausal transition.

This study provides initial evidence that L1CAM-enriched EV miRNA profiles differ between early and late perimenopause and that these differences map onto brain-relevant signaling networks implicated in hormone-sensitive neuronal adaptation. In this pilot sample, nEV miRNA levels did not track a single estradiol measure, suggesting they may not simply reflect acute endocrine state at the time of sampling. They may instead be more sensitive to longer-timescale neuroendocrine dynamics across the transition (e.g., variability and cumulative exposure), which should be tested in longitudinal designs with repeated hormone sampling. These findings support longitudinal investigation of nEV miRNAs as potential biomarkers of brain aging and neurobiological vulnerability during the midlife menopausal transition.

## Data Availability

All data produced in the present study are available upon reasonable request to the authors

