## Supplemental Figures for "Neuron-Enriched Extracellular Vesicle MicroRNAs Reflect Hormone-Sensitive Neural Pathway Changes from Early to Late Perimenopause"

### Supplement

**Supplementary Table S1.** Directional concordance between pooled discovery screening and individual-level qPCR validation of nEV miRNAs across the menopausal transition.

| miRNA | Discovery log2FC | Discovery direction | Validation log2FC | Validation direction | Concordant |
|---|---|---|---|---|---|
| miR-24-3p | -15.94 | Down in Late | -0.26 | Down in Late | Yes |
| miR-30b-5p | -14.77 | Down in Late | -0.33 | Down in Late | Yes |
| miR-150-5p | -10.65 | Down in Late | -0.65 | Down in Late | Yes |
| miR-152-3p | -8.09 | Down in Late | -0.39 | Down in Late | Yes |
| miR-197-3p | -7.47 | Down in Late | -0.97 | Down in Late | Yes |
| miR-194-5p | -7.12 | Down in Late | -0.59 | Down in Late | Yes |
| miR-425-3p | -6.87 | Down in Late | -0.35 | Down in Late | Yes |
| miR-22-5p | -6.35 | Down in Late | 0.35 | Up in Late | No |
| miR-584-5p | -6.15 | Down in Late | -0.2 | Down in Late | Yes |
| miR-30a-5p | -6.1 | Down in Late | -1.13 | Down in Late | Yes |

**Supplementary Table S2. Type III Tests of Fixed Effects**
**Primary mixed model:** ΔCq ~ miRNA Species + STRAW Stage + miRNA Species x STRAW Stage

| Fixed Effect | Num df | Den df | F | p | partial $\eta^2$ |
|---|---|---|---|---|---|
| miRNA Species | 9 | 69.48 | 530.77 | < .001 | 0.99 |
| STRAW Stage | 1 | 57.58 | 5.57 | .022 | 0.09 |
| miRNA Species x STRAW Stage | 9 | 69.48 | 1.27 | .270 | 0.14 |

**Supplementary Table S3. Type III Tests of Fixed Effects**
**Age-adjusted mixed model:** ΔCq ~ miRNA Species + STRAW Stage + miRNA Species x STRAW Stage + Age.

| Fixed Effect | Num df | Den df | F | p | partial $\eta^2$ |
|---|---|---|---|---|---|
| miRNA Species | 9 | 69.30 | 529.82 | < .001 | 0.99 |
| STRAW Stage | 1 | 54.67 | 5.48 | .023 | 0.09 |
| miRNA Species × STRAW Stage | 9 | 69.30 | 1.27 | .270 | 0.14 |
| Age | 1 | 38.37 | 0.08 | .786 | < 0.01 |

**Supplementary Table S4. Type III Tests of Fixed Effects**
**Greene symptom–adjusted model:** ΔCq ~ miRNA Species + Greene Climacteric Scale Total Score.

| Fixed Effect | Num df | Den df | F | p | partial $\eta^2$ |
|---|---|---|---|---|---|
| miRNA Species | 9 | 73.88 | 539.87 | < .001 | 0.99 |
| Greene Total | 1 | 38.86 | 1.60 | .214 | 0.04 |

**Supplementary Table S5. Type III Tests of Fixed Effects**
**Estradiol-adjusted model:** ΔCq ~ miRNA Species + Estradiol (log).

| Fixed Effect | Num df | Den df | F | p | partial $\eta^2$ |
|---|---|---|---|---|---|
| miRNA Species | 9 | 71.47 | 536.96 | < .001 | 0.99 |

| Fixed Effect | Num df | Den df | F | p | partial $\eta^2$ |
|---|---|---|---|---|---|
| Estradiol (log) | 1 | 38.81 | 0.00 | .950 | < 0.01 |

**Supplementary Table S6. Estimated STRAW Stage Effect (Late − Early).**

| Effect | Estimate (ΔCq) | SE | 95% CI | p | FC ($2^{-\Delta\Delta Cq}$) |
|---|---|---|---|---|---|
| STRAW Stage | 0.51 | 0.21 | [0.08, 0.93] | .022 | 0.70 |

**Supplementary Table S7. Estimated STRAW Stage Effect (Late − Early) for the Age-Adjusted Model.**

| Effect | Estimate (ΔCq) | SE | 95% CI | p | FC ($2^{-\Delta\Delta Cq}$) |
|---|---|---|---|---|---|
| STRAW Stage | 0.52 | 0.22 | [0.08, 0.96] | .023 | 0.69 |

**Supplementary Table S8. Estimated Effect of Greene Climacteric Scale (Total Score).**

| Effect | Estimate (ΔCq per SD) | SE | 95% CI | p |
|---|---|---|---|---|
| Greene Total | 0.13 | 0.10 | [−0.08, 0.33] | .214 |

**Supplementary Table S9. Estimated Effect of Estradiol (Log).**

| Effect | Estimate (ΔCq per SD) | SE | 95% CI | p |
|---|---|---|---|---|
| Estradiol (log) | 0.01 | 0.10 | [−0.20, 0.22] | .950 |

**Supplementary Tables S10.** PANTHER Pathway Enrichment Analysis.

PANTHER Pathways (Top 10 results)

| panther_pathway | Total | Observed | Expected | OverRep_UnderRepresented | FoldEnrich | bonf_p |
|---|---|---|---|---|---|---|
| Gonadotropin-releasing hormone receptor pathway (P06664) | 231 | 108 | 60.01 | + | 1.80 | 0.00e+00 |
| PDGF signaling pathway (P00047) | 144 | 75 | 37.41 | + | 2.01 | 0.00e+00 |
| Integrin signalling pathway (P00034) | 192 | 88 | 49.88 | + | 1.76 | 5.00e-07 |
| EGF receptor signaling pathway (P00018) | 134 | 65 | 34.81 | + | 1.87 | 3.80e-06 |
| Angiogenesis (P00005) | 169 | 74 | 43.90 | + | 1.69 | 8.21e-05 |
| FGF signaling pathway (P00021) | 122 | 56 | 31.69 | + | 1.77 | 3.12e-04 |
| Interferon-gamma signaling pathway (P00035) | 30 | 18 | 7.79 | + | 2.31 | 1.26e-02 |
| CCKR signaling map (P06959) | 173 | 68 | 44.94 | + | 1.51 | 1.71e-02 |
| p53 pathway (P00059) | 88 | 39 | 22.86 | + | 1.71 | 3.08e-02 |
| PI3 kinase pathway (P00048) | 55 | 27 | 14.29 | + | 1.89 | 4.16e-02 |

**Supplementary Tables S11-S13.** Gene Ontology Enrichment for Biological Process, Molecular Function, and Cellular Components.

**Supplementary Table S11**

**Gene Ontology - Biological Process (Top 20 Results)**

- NB: These results are for GO categories larger than 10 members and smaller than 1500 members. The rationale for limiting GO categories by size is that the categories of extreme size (either very large or very small) are vague (i.e., difficult to interpret) and the statistical parameters are less trustworthy compared to average-sized GO categories.

| BioProcess | Total | Observed | Expected | OverRep_UnderRepresented | FoldEnrich | bonf_p |
|---|---|---|---|---|---|---|
| detection of chemical stimulus involved in sensory perception (GO:0050907) | 485 | 5 | 125.99 | - | .04 | 0 |
| detection of chemical stimulus involved in sensory perception of smell (GO:0050911) | 437 | 4 | 113.52 | - | .04 | 0 |

| BioProcess | Total | Observed | Expected | OverRep_UnderRepresented | FoldEnrich | bonf_p |
|---|---|---|---|---|---|---|
| detection of chemical stimulus (GO:0009593) | 522 | 13 | 135.60 | - | .10 | 0 |
| sensory perception of chemical stimulus (GO:0007606) | 550 | 19 | 142.87 | - | .13 | 0 |
| sensory perception of smell (GO:0007608) | 469 | 12 | 121.83 | - | .10 | 0 |
| detection of stimulus involved in sensory perception (GO:0050906) | 563 | 34 | 146.25 | - | .23 | 0 |
| neurogenesis (GO:0022008) | 1433 | 576 | 372.25 | + | 1.55 | 0 |
| generation of neurons (GO:0048699) | 1235 | 504 | 320.81 | + | 1.57 | 0 |
| neuron differentiation (GO:0030182) | 1156 | 474 | 300.29 | + | 1.58 | 0 |
| vesicle-mediated transport (GO:0016192) | 1321 | 527 | 343.15 | + | 1.54 | 0 |
| detection of stimulus (GO:0051606) | 694 | 63 | 180.28 | - | .35 | 0 |
| negative regulation of signal transduction (GO:0009968) | 1423 | 539 | 369.65 | + | 1.46 | 0 |
| positive regulation of transcription by RNA polymerase II (GO:0045944) | 1262 | 488 | 327.83 | + | 1.49 | 0 |
| regulation of multicellular organismal development (GO:2000026) | 1419 | 535 | 368.61 | + | 1.45 | 0 |
| neuron development (GO:0048666) | 916 | 374 | 237.95 | + | 1.57 | 0 |
| anatomical structure formation involved in morphogenesis (GO:0048646) | 1001 | 397 | 260.03 | + | 1.53 | 0 |
| regulation of cell migration (GO:0030334) | 957 | 381 | 248.60 | + | 1.53 | 0 |
| regulation of locomotion (GO:0040012) | 1060 | 413 | 275.35 | + | 1.50 | 0 |
| tube development (GO:0035295) | 936 | 373 | 243.14 | + | 1.53 | 0 |
| protein transport (GO:0015031) | 1110 | 427 | 288.34 | + | 1.48 | 0 |

**Supplementary Table S12**

**Gene Ontology - Molecular Function (Top 20 Results)**

- NB: These results are for GO categories larger than 10 members and smaller than 1500 members. The rationale for limiting GO categories by size is that the categories of extreme size (either very large or very small) are vague (i.e., difficult to interpret) and the statistical parameters are less trustworthy compared to average-sized GO categories.

| MolFunction | Total | Observed | Expected | OverRep_UnderRepresented | FoldEnrich | bonf_p |
|---|---|---|---|---|---|---|
| olfactory receptor activity (GO:0004984) | 435 | 3 | 113.00 | - | .03 | 0e+00 |
| G protein-coupled receptor activity (GO:0004930) | 879 | 109 | 228.33 | - | .48 | 0e+00 |
| molecular adaptor activity (GO:0060090) | 1441 | 519 | 374.32 | + | 1.39 | 0e+00 |
| phosphotransferase activity, alcohol group as acceptor (GO:0016773) | 682 | 277 | 177.16 | + | 1.56 | 0e+00 |
| protein-macromolecule adaptor activity (GO:0030674) | 1301 | 471 | 337.96 | + | 1.39 | 0e+00 |
| kinase activity (GO:0016301) | 731 | 291 | 189.89 | + | 1.53 | 0e+00 |
| transcription cis-regulatory region binding (GO:0000976) | 1440 | 506 | 374.06 | + | 1.35 | 0e+00 |
| transcription regulatory region nucleic acid binding (GO:0001067) | 1441 | 506 | 374.32 | + | 1.35 | 0e+00 |
| odorant binding (GO:0005549) | 119 | 1 | 30.91 | - | .03 | 0e+00 |
| RNA polymerase II transcription regulatory region sequence-specific DNA binding (GO:0000977) | 1341 | 470 | 348.35 | + | 1.35 | 0e+00 |
| transferase activity, transferring phosphorus-containing groups (GO:0016772) | 901 | 335 | 234.05 | + | 1.43 | 0e+00 |
| cis-regulatory region sequence-specific DNA binding (GO:0000987) | 1146 | 410 | 297.69 | + | 1.38 | 0e+00 |
| RNA polymerase II cis-regulatory region sequence-specific DNA binding (GO:0000978) | 1122 | 402 | 291.46 | + | 1.38 | 0e+00 |
| cytoskeletal protein binding (GO:0008092) | 1008 | 367 | 261.84 | + | 1.40 | 0e+00 |

| MolFunction | Total | Observed | Expected | OverRep_UnderRepresented | FoldEnrich | bonf_p |
|---|---|---|---|---|---|---|
| ATP binding (GO:0005524) | 1478 | 502 | 383.94 | + | 1.31 | 0e+00 |
| protein kinase activity (GO:0004672) | 572 | 225 | 148.59 | + | 1.51 | 0e+00 |
| kinase binding (GO:0019900) | 782 | 291 | 203.14 | + | 1.43 | 0e+00 |
| protein serine/threonine kinase activity (GO:0004674) | 435 | 179 | 113.00 | + | 1.58 | 0e+00 |
| protein-containing complex binding (GO:0044877) | 1299 | 446 | 337.44 | + | 1.32 | 0e+00 |
| DNA-binding transcription factor activity, RNA polymerase II-specific (GO:0000981) | 1332 | 451 | 346.01 | + | 1.30 | 1e-07 |

**Supplementary Table S13.**

**Gene Ontology - Cellular Components (Top 20 Results)**

- NB: These results are for GO categories larger than 10 members and smaller than 1500 members. The rationale for limiting GO categories by size is that the categories of extreme size (either very large or very small) are vague (i.e., difficult to interpret) and the statistical parameters are less trustworthy compared to average-sized GO categories.

| CellComponents | Total | Observed | Expected | OverRep_UnderRepresented | FoldEnrich | bonf_p |
|---|---|---|---|---|---|---|
| glutamatergic synapse (GO:0098978) | 585 | 285 | 151.96 | + | 1.88 | 0 |
| postsynapse (GO:0098794) | 782 | 343 | 203.14 | + | 1.69 | 0 |
| neuron projection (GO:0043005) | 1358 | 526 | 352.76 | + | 1.49 | 0 |
| immunoglobulin complex (GO:0019814) | 198 | 1 | 51.43 | - | .02 | 0 |
| axon (GO:0030424) | 664 | 287 | 172.49 | + | 1.66 | 0 |
| presynapse (GO:0098793) | 647 | 280 | 168.07 | + | 1.67 | 0 |
| somatodendritic compartment (GO:0036477) | 857 | 345 | 222.62 | + | 1.55 | 0 |
| dendrite (GO:0030425) | 630 | 266 | 163.65 | + | 1.63 | 0 |
| dendritic tree (GO:0097447) | 632 | 266 | 164.17 | + | 1.62 | 0 |
| neuron to neuron synapse (GO:0098984) | 403 | 187 | 104.69 | + | 1.79 | 0 |
| chromatin (GO:0000785) | 1399 | 509 | 363.41 | + | 1.40 | 0 |
| Golgi membrane (GO:0000139) | 707 | 290 | 183.66 | + | 1.58 | 0 |

| CellComponents | Total | Observed | Expected | OverRep_UnderRepresented | FoldEnrich | bonf_p |
|---|---|---|---|---|---|---|
| cytoplasmic vesicle membrane (GO:0030659) | 1273 | 469 | 330.68 | + | 1.42 | 0 |
| plasma membrane region (GO:0098590) | 1336 | 486 | 347.05 | + | 1.40 | 0 |
| vesicle membrane (GO:0012506) | 1294 | 473 | 336.14 | + | 1.41 | 0 |
| synaptic membrane (GO:0097060) | 451 | 200 | 117.15 | + | 1.71 | 0 |
| asymmetric synapse (GO:0032279) | 367 | 169 | 95.33 | + | 1.77 | 0 |
| postsynaptic specialization (GO:0099572) | 387 | 175 | 100.53 | + | 1.74 | 0 |
| postsynaptic density (GO:0014069) | 351 | 161 | 91.18 | + | 1.77 | 0 |
| anchoring junction (GO:0070161) | 918 | 346 | 238.47 | + | 1.45 | 0 |

**Supplementary Table S14.**

**Global Results (Top 20 Results; Arranged by P-Values)**

- NB: These results are for GO categories larger than 10 members and smaller than 1500 members. The rationale for limiting GO categories by size is that the categories of extreme size (either very large or very small) are vague (i.e., difficult to interpret) and the statistical parameters are less trustworthy compared to average-sized GO categories.

| GO_category | Total | Observed | Expected | OverRep_UnderRepresented | FoldEnrich | bonf_p |
|---|---|---|---|---|---|---|
| detection of chemical stimulus involved in sensory perception (GO:0050907) | 485 | 5 | 125.99 | - | .04 | 0 |
| olfactory receptor activity (GO:0004984) | 435 | 3 | 113.00 | - | .03 | 0 |
| detection of chemical stimulus involved in sensory perception of smell (GO:0050911) | 437 | 4 | 113.52 | - | .04 | 0 |
| detection of chemical stimulus (GO:0009593) | 522 | 13 | 135.60 | - | .10 | 0 |

| GO_category | Total | Observed | Expected | OverRep_UnderRepresented | FoldEnrich | bonf_p |
|---|---|---|---|---|---|---|
| sensory perception of chemical stimulus (GO:0007606) | 550 | 19 | 142.87 | - | .13 | 0 |
| sensory perception of smell (GO:0007608) | 469 | 12 | 121.83 | - | .10 | 0 |
| detection of stimulus involved in sensory perception (GO:0050906) | 563 | 34 | 146.25 | - | .23 | 0 |
| neurogenesis (GO:0022008) | 1433 | 576 | 372.25 | + | 1.55 | 0 |
| glutamatergic synapse (GO:0098978) | 585 | 285 | 151.96 | + | 1.88 | 0 |
| generation of neurons (GO:0048699) | 1235 | 504 | 320.81 | + | 1.57 | 0 |
| neuron differentiation (GO:0030182) | 1156 | 474 | 300.29 | + | 1.58 | 0 |
| vesicle-mediated transport (GO:0016192) | 1321 | 527 | 343.15 | + | 1.54 | 0 |
| detection of stimulus (GO:0051606) | 694 | 63 | 180.28 | - | .35 | 0 |
| postsynapse (GO:0098794) | 782 | 343 | 203.14 | + | 1.69 | 0 |
| neuron projection (GO:0043005) | 1358 | 526 | 352.76 | + | 1.49 | 0 |
| immunoglobulin complex (GO:0019814) | 198 | 1 | 51.43 | - | .02 | 0 |
| negative regulation of signal transduction (GO:0009968) | 1423 | 539 | 369.65 | + | 1.46 | 0 |
| positive regulation of transcription by RNA | 1262 | 488 | 327.83 | + | 1.49 | 0 |

| GO_category | Total | Observed | Expected | OverRep_UnderRepresented | FoldEnrich | bonf_p |
|---|---|---|---|---|---|---|
| polymerase II (GO:0045944) | | | | | | |
| G protein-coupled receptor activity (GO:0004930) | 879 | 109 | 228.33 | - | .48 | 0 |
| regulation of multicellular organismal development (GO:2000026) | 1419 | 535 | 368.61 | + | 1.45 | 0 |

**Supplementary Table S15.**

**Global Results (Top 20 Results; Arranged by Effect Size)**

- NB: These results are for GO categories larger than 10 members and smaller than 1500 members. The rationale for limiting GO categories by size is that the categories of extreme size (either very large or very small) are vague (i.e., difficult to interpret) and the statistical parameters are less trustworthy compared to average-sized GO categories.

| GO_category | Total | Observed | Expected | OverRep_UnderRepresented | FoldEnrich | bonf_p |
|---|---|---|---|---|---|---|
| N6-methyladenosine-containing RNA reader activity (GO:1990247) | 12 | 10 | 3.12 | + | 3.21 | 0.16900 |
| phosphatidylinositol monophosphate phosphatase activity (GO:0052744) | 16 | 13 | 4.16 | + | 3.13 | 0.01870 |
| commissural neuron axon guidance (GO:0071679) | 14 | 11 | 3.64 | + | 3.02 | 0.54100 |
| phosphatidylinositol-3-phosphate phosphatase activity (GO:0004438) | 14 | 11 | 3.64 | + | 3.02 | 0.18300 |
| neuronal ion channel clustering (GO:0045161) | 13 | 10 | 3.38 | + | 2.96 | 1.00000 |
| negative regulation of nitric oxide biosynthetic process (GO:0045019) | 13 | 10 | 3.38 | + | 2.96 | 1.00000 |
| regulation of calcium ion import across plasma membrane (GO:1905664) | 13 | 10 | 3.38 | + | 2.96 | 1.00000 |

| GO_category | Total | Observed | Expected | OverRep_UnderRepresented | FoldEnrich | bonf_p |
|---|---|---|---|---|---|---|
| negative regulation of nitric oxide metabolic process (GO:1904406) | 13 | 10 | 3.38 | + | 2.96 | 1.00000 |
| negative regulation of sprouting angiogenesis (GO:1903671) | 13 | 10 | 3.38 | + | 2.96 | 1.00000 |
| reelin-mediated signaling pathway (GO:0038026) | 12 | 9 | 3.12 | + | 2.89 | 1.00000 |
| regulation of stem cell division (GO:2000035) | 12 | 9 | 3.12 | + | 2.89 | 1.00000 |
| neurotransmitter receptor transport to postsynaptic membrane (GO:0098969) | 12 | 9 | 3.12 | + | 2.89 | 1.00000 |
| regulation of stress granule assembly (GO:0062028) | 12 | 9 | 3.12 | + | 2.89 | 1.00000 |
| prostate gland growth (GO:0060736) | 12 | 9 | 3.12 | + | 2.89 | 1.00000 |
| ATP-dependent chromatin remodeler activity (GO:0140658) | 24 | 18 | 6.23 | + | 2.89 | 0.00223 |
| phosphatidylinositol-3,5-bisphosphate 3-phosphatase activity (GO:0052629) | 12 | 9 | 3.12 | + | 2.89 | 1.00000 |
| dystroglycan binding (GO:0002162) | 12 | 9 | 3.12 | + | 2.89 | 1.00000 |
| epithelial cell fate commitment (GO:0072148) | 15 | 11 | 3.90 | + | 2.82 | 1.00000 |
| negative regulation of axon regeneration (GO:0048681) | 15 | 11 | 3.90 | + | 2.82 | 1.00000 |
| facial nerve morphogenesis (GO:0021610) | 11 | 8 | 2.86 | + | 2.80 | 1.00000 |
